# Thromboembolic events and major bleeding with warfarin, non-vitamin K antagonist oral anticoagulants or antiplatelets following mitral valve repair: A systematic review and meta-analysis

**DOI:** 10.1101/2022.10.29.22281662

**Authors:** Bilal H Kirmani, Sion G Jones, Andrew D Muir, Graham S. Hillis, Ralph Stewart, Cheng-Hon Yap, Rebecca H Maier, Helen C Hancock, James Wason, Benoy N Shah, Richard Whitlock, Gregory Y.H. Lip, Enoch Akowuah

**Author notes:** Corresponding author Bilal Kirmani, Department of Cardiac Surgery, Liverpool Heart and Chest, Hospital, Thomas Drive, Liverpool, L14 3PE, United Kingdom.

## Abstract

**Background:** International guidelines recommend the use of a vitamin K antagonist (VKA, e.g. warfarin) for the first three months after mitral repair as a Class IIa recommendation with Level of Evidence C. High rates of atrial fibrillation, thrombogenicity of the non-endothelialised repair components, and high rates of antiplatelet resistance are cited as rationale. However, surveys of practice indicate that surgeon compliance is low, suggesting uncertainty of the evidence. We sought to establish the best current evidence by conducting a systematic review as a precursor to a randomised trial.

**Methods:** We included prospective randomised and retrospective observational studies of adult patients undergoing mitral valve repair with no other indications for anticoagulation. Studies had to compare a VKA to a non-vitamin K antagonist oral anticoagulants (NOAC), anti-platelet or placebo/null treatment (“Other”). Outcomes had to include thromboembolic complications, major bleeding or mortality within three months. The Cochrane Register, Medline, Embase and Clinical Trials Registries were checked. Risk of Bias assessments were conducted using the ROBINS-I tool as part of the GRADE Pro methodology. Quantitative synthesis was agreed following review of the methodology and a random effects model using the Mantel-Haenszel method employed.

**Results:** There were no randomised controlled trials comparing VKA to other treatment strategies. Six observational studies comprising 5291 patients (2925 receiving VKA, 807 antiplatelet, 104 NOAC and 1455 null treatment) were included. Four studies reporting early thromboembolic risk found that the use of a VKA may not be associated with a reduction in risk (relative risk [RR] 0.54, 95% CI 0.13 – 2.35, p=0.41, I^2^ = 68%, overall certainty of evidence = very low). Three studies reporting late thromboembolism also showed that use of VKA may not be associated with risk reduction (RR 0.84, 95% CI 0.57 – 1.24, p=0.37, I^2^ = 23%).

Two studies reported on early risks of major bleeding and found VKA treatment had very uncertain effects (RR 0.60, 95% CI 0.36 – 1.02, p=0.06, I^2^=0%, certainty of evidence = very low). Late bleeding reported by four studies was uncertain but seemed to show no risk reduction with VKA (RR 0.97, 95% CI 0.37 – 2.52, p=0.95, I^2^=73%). Three studies found an unclear effect of VKA use on early mortality (RR 0.39, 95% CI 0.10 – 1.51, p=0.17, I^2^=35%, certainty of evidence = low). Two studies comparing VKA with antiplatelets and no thromboprophylaxis respectively, showed there may be risk reduction in late mortality (RR 0.72, 95% CI 0.59 – 0.87, p=0.0006, I^2^=0%, certainty of evidence = very low), although this effect was weighted by a single study.

**Conclusion:** There is insufficient good quality evidence to inform anticoagulation, anti-platelet or null treatment practices following mitral valve repair. A high-quality trial is therefore required in this setting, with particular methodological focus on aspects found to increase risk of bias in current studies.

**Funding:** There was no funding for this systematic review.

**Registration:** The review protocol was published in PROSPERO (CRD42021271198).

## Introduction

Mitral valve prolapse is common, affecting 2-3% of the population and an estimated 176 million people worldwide (1). Prevalence is highest in countries with high socio-demographic indices (SDI) such as North America and Europe, but this may reflect better access services. The definitive treatment for degenerative mitral valve diseases is surgical intervention, with repair preferred due to low peri-operative mortality, morbidity and improved long term durability (2,3).

While unadjusted operative mortality following mitral valve surgery for degenerative disease is 0.8% for mitral valve repair (compared to 3.1% for mitral valve replacement) the incidence of thromboembolic events following surgery resulting in end-organ ischemia is higher (4). Within 30 days of surgery, ischaemic stroke rates are up to 2.1%, with a further 1.5% event rate between 30 and 180 days. Annual incidence thereafter falls to 0.9% (5,6).

To mitigate these risks, some surgeons use anticoagulation or anti-platelet treatment following mitral valve repair anecdotally to allow the thrombogenic sewing ring to endothelialise. Prophylaxis for the period of highest post-operative atrial fibrillation risk is also cited as the rationale, although this logic is not extended to all cardiac surgical procedures where atrial fibrillation may occur. From international surveys of practice (pending publication), the choice of treatment modality and duration are subject to wide variation. The 2021 European Society of Cardiology (ESC) / European Association for Cardio-Thoracic Surgery (EACTS) guidelines for the management of valvular heart disease suggest that oral anticoagulation with a Vitamin K Antagonist (VKA, eg. warfarin) should be considered for the first three months following mitral valve repair in the absence of atrial fibrillation (2). This is a Class IIa recommendation with Level C evidence without supporting references, however compliance with this guideline is approximately 30% from a recent survey of UK surgeons (pending publication). A further third of surgeons will employ anticoagulation with non-vitamin K antagonist oral anticoagulants (NOACs, also known as direct oral anticoagulants or DOACS) for the same period, with the final third of surgeons using antiplatelet medications either for 3 months or lifelong. By contrast, the 2020 American College of Cardiology (ACC) / American Heart Association (AHA) guidelines do not make any recommendations on anticoagulation or antiplatelet management following mitral valve repair (7).

A systematic review from 2006 (8) was used to inform the EACTS guideline on antiplatelet and anticoagulation management in cardiac surgery (9). This was published two years before the first NOAC, dabigatran, was first licensed by the European Medicines Agency, and therefore could not account for that modality. The data summarized were not of high quality: of the 12 papers included, 9 were retrospective observational cohort studies. None randomised patients between warfarin and an antiplatelet medication, and all included a heterogenous group of repair and tissue replacement patients. There are, therefore, no meta-analyses of existing literature to support recommendations or practice and the National Institute for Health and Care Excellence (NICE) in the United Kingdom has identified this as an area requiring research (10). Accordingly, we undertook this systematic review and meta-analysis as a precursor to an international randomised controlled trial to answer the clinical question “In patients undergoing mitral valve repair, how does anticoagulation with warfarin compare with either novel oral anticoagulants or anti-platelets in the prevention of thromboembolic or major bleeding complications or mortality”.

## Methods

We undertook a systematic review and meta-analysis according to the PRISMA 2020 guidelines (11). The review was registered on PROSPERO (CRD42021271198).

### Eligibility criteria

We included all comparative studies with the following PICOS characteristics: (i) Population - adult patients undergoing mitral valve repair and with no other indications for anticoagulation; (ii) Intervention –antiplatelet, NOAC or placebo/null treatment following surgery; (iii) Control – short term (6-12 week) warfarin treatment following surgery; (iv) Outcomes – at 3 months and longest available follow-up [Thromboembolic complications (including cerebrovascular accident, transient ischemic attack, mesenteric ischaemia, limb ischemia, or intracardiac thrombus); Major bleeding complications (as defined by International Society of Thrombosis and Hemostasis (12)); and All-cause mortality]; (v) Study type – prospective randomised controlled trials, comparative observational retrospective cohorts and propensity matched studies; and (vi) Setting – cardiac surgery centres worldwide

We did not exclude studies published in languages other than English, as long as an English translation of the title was available for screening.

### Information Sources

We conducted searches for eligible studies within each of the following databases: Cochrane Central Register of Controlled Trials (CENTRAL) (1992 – 21 July 2021); MEDLINE (via the PubMed interface) (1946 – 21 July 2021); Embase (via the Ovid interface) (1980 – 21 July 2021); Clinical trials registries (US National Institutes of Health Ongoing Trials Register at ClinicalTrials.gov; World Health Organization International Clinical Trials Registry Platform at apps.who.int/trialsearch/)

Search strategies for all databases are presented in Appendix I. In addition, we searched the reference lists of all eligible reports and undertook forward citation tracking using the PubMed interface.

### Selection process

Following database searching, all identified studies were imported into Zotero reference manager. Duplicates were assessed and removed manually. Two reviewers (BHK and SGJ) then independently screened all titles and abstracts for eligibility and inclusion with a third author (ADM) arbitrating disagreements.

### Data collection process

Two independent reviewers (BHK and SGJ), each blinded to the others’ findings, extracted data from studies directly into standardised electronic spreadsheets, discussing any discrepancies and with arbitration by a third reviewer (ADM) in the event of disagreement in the interpretation of data. Data were then transferred by BHK into Review Manager 5 software (Cochrane Collaboration, 2014) (13) for synthesis where appropriate.

### Data Items

As much relevant data to help understand heterogeneity in studies were collected as possible, as well the study outcomes. The following list was employed to guide the data collection proforma, but variables not cited that were otherwise considered of importance to the review process were recorded in separate notes: Study design (e.g. prospective, retrospective, randomised, propensity matched, case-control, cohort etc); PICOS details as described above; Study location(s); Study funding; Conflict of Interests; Subjective risks of bias, uncertainty or confounders; Total number of patients, and patients in each arm where relevant; Demographics and baseline characteristics (age, gender, pathology, surgery, cardiopulmonary bypass times and cross-clamp times); Definitions and timings of outcome measures, if given. Where serial measures were taken, early (3 months) and late (longest follow-up) were recorded; Number of events / measure of effect (as median and interquartile range or mean and standard deviation, with Cochrane corrections if data presented as standard error).

### Risk of Bias Assessment

The Risk Of Bias In Non-Randomized Studies - of Interventions (ROBINS-I) tool from the Cochrane Bias Methods Group (14) was used to assess and report bias and quality. The hypothetical pragmatic “target” trial for the study was specified as an individually randomized trial of adult patients undergoing mitral valve repair including those having concomitant surgery. The control group was specified as vitamin K antagonist (VKA) anticoagulation for 6-12 weeks following surgery. The intervention group was specified as anticoagulation with NOACs or antiplatelet agents or placebo/null treatment. The domains considered to be potential sources of bias included:

- Confounding – risk of variables affecting both treatment allocation and outcomes
- Selection – risk of selection bias with inclusion or exclusion criteria
- Classification – risk of classification errors of treatment arm
- Deviations from planned interventions – crossovers or withdrawals
- Missing data – incomplete or missing data
- Measurements – measurement biases including those affected by absence of allocation concealment
- Selective reporting – biases of reporting limited outcomes

Two independent reviewers (BHK and SGJ) assessed quality by these criteria, with discussion to resolve any differences, and a third reviewer (ADM) provided arbitration in the event of a disagreement. Inconsistency, imprecision and indirectness were also assessed to provide overall certainty levels for the evidence, using GRADEPro to summarise findings and inform data synthesis.

### Effect Measures

As all outcomes measured were dichotomous variables, risk ratios were used as the effect measure as these are usually the easiest to interpret.

### Synthesis methods

Studies were eligible for synthesis according to informal analysis of the methodology. Review authors had to agree that the studies represented accepted practice for management of post-operative anticoagulation or anti-platelet treatment with representative patient cohorts. Within each outcome, studies were sub-grouped according to the choice of non-VKA management (i.e. NOAC, antiplatelet or no treatment). As all outcomes were dichotomous, no data conversions were required. Missing data were explored by communication with the study authors.

Data were presented for individual studies in tabulated format in the description of included studies (Table 1) with Summary of Findings reporting the synthesised data (Table 2). Individual study risk of bias, as determined by the ROBINS-I tool, was shown graphically in Figure 1. Graphical representation of each of the outcomes was shown with forest plots (Figures 2-7), subgrouping according to non-VKA strategy and ordering studies by date.

**Table 1.** Characteristics of included studies

| Study | Design | Participants | Interventions | Outcomes | Comments |
| --- | --- | --- | --- | --- | --- |
| Meurin (2008) [15] | Prospective multicentre non-randomised | 350 consecutive patients following mitral valve repair (including other procedures). Subgroup analysis to exclude AF and concomitant surgery included 185 patients and was included. | 1. VKA<br>2. VKA+ASA<br>3. ASA<br>4. No prophylaxis | FOLLOWUP<br>6 weeks<br>THROMBOEMBOLIC<br>Thromboembolic events<br>BLEEDING<br>Hemorrhagic events<br>OTHER<br>Arrhythmia, infection, signs of heart failure, and occurrence or worsening of mitral regurgitation murmur | 1. Main analysis includes pre-op AF and concomitant surgery<br>2. Subgroup analysis excludes pre- AND post-op AF, and concomitant surgery |
| Noohi (2020) [16] | Mixed cohort study (prospective patients for rivaroxaban and retrospective cohort of propensity matched patients for | 297 patients undergoing mitral valve repair (including those with concomitant surgery)<br>Excluded: redo surgery, severe renal impairment, other reasons for anticoagulation (except AF), those over 75y | 1. VKA (warfarin)<br>2. DOAC (rivaroxaban) | FOLLOW UP<br>1 year<br>THROMBOEMBOLIC<br>Ischemic stroke or transient ischemic attack<br>BLEEDING<br>major or clinically relevant non-major bleeding according to the classification of the | Excluded patients requiring reoperation in the post operative period. Excluded those who died in the post-operative period. Anticoagulation continued in 11-18% of patients beyond 3 months, including those with pre-operative AF. |
|  | warfarin) - effectively a propensity matched study |  |  | International Society on Thrombosis and Haemostasis (ISTH) |  |
| Paparella (2016) [17] | Retrospective multicentre international cohort study | 1882 patients<br>Inclusion Criteria<br>Included patients were aged >18 years at time of surgery underwent isolated MVR with mitral ring implantation to treat mitral regurgitation, and in sinus rhythm at hospital discharge.<br>Exclusion Criteria<br>Exclusion criteria included any history of atrial fibrillation, any combined surgical procedure associated with mitral valve repair, and treatment with both oral anticoagulation and antiplatelet (80 or 100 mg once daily). | 1. VKA<br>2. aspirin | FOLLOW UP<br>6 months<br>THROMBOEMBOLIC<br>An arterial event was defined as the presence of 1 of the following within 6 months of the mitral valve operation: cardiac valvular or mural thrombus confirmed by echocardiogram, intracranial event confirmed by computed tomography/magnetic resonance imaging, transient ischemic attack, or limb or mesenteric thromboembolic event confirmed by arteriogram, magnetic resonance imaging, or computed tomography.<br>BLEEDING<br>Major bleeding within the first 48 hours after surgery was defined per the Bleeding Academic Research | Excluded patients going into AF post-operatively |
|  |  |  |  | Consortium: perioperative intracranial bleeding within 48 hours, and/or reoperation after closure of sternotomy for the purpose of controlling bleeding, and/or transfusion of >5 units whole blood or packed red blood cells within a 48-hour period (cell saver products were not counted), and/or chest tube output >2 L within a 24-hour period.<br>Major bleeding that occurred at least 48 hours after surgery was defined per the International Society of Thrombosis and Haemostasis definition<sup>8</sup>: fatal bleeding; and/or symptomatic bleeding in a critical area or organ, such as intracranial, intraspinal, intraocular, retroperitoneal, intra-articular or pericardial, or intramuscular with compartment syndrome; and/or bleeding causing a fall in hemoglobin level of 3.0 g/dL or more or leading to transfusion of 2 or more units<br>of whole blood or red cells. |  |
| Valeur<br>(2016) [18] | Retrospective cohort study | 2188 patients from Danish registry<br>all patients without previous cardiac surgery having mitral valve repair surgery performed in the period between 1 January 1997 and 31 December 2012 (including those having concomitant surgery)<br>Excluded patients on non-VKA anticoagulation | 1. VKA (warfarin)<br>2. Non-VKA (including aspirin and no prophylaxis) | FOLLOW UP<br>Median 4.9 years<br>(analyses starting 7 days after discharge)<br>PRIMARY<br>Composite of death or stroke<br>SECONDARY<br>Mortality, stroke or major bleeding<br>THROMBOEMBOLIC<br>not specified<br>BLEEDING<br>major bleeding requiring hospital admission with gastrointestinal bleeding/ulcer, intracranial and other serious bleeding as airway bleeding or bleeding from the urinary tract |  |
| Van der Wall<br>(2018) [19] | Retrospective observational multicentre cohort study of consecutive adult patients who | 469 patients<br>First MVr with or without concomitant tricuspid valve repair (TVr) between 2004 and 2016<br>Excluded other concomitant cardiac procedures than TVr, had previous cardiac surgery or were diagnosed with AF preoperatively | 1. VKA for 6-12 weeks (then either continued or replaced by aspirin)<br>2. Aspirin 80mg | FOLLOW UP<br>3 months<br>PRIMARY<br>Composite of thromboembolic and major bleeding complications<br>THROMBOEMBOLIC<br>criteria from International |  |
|  | underwent mitral valve repair |  | lifelong | Society on Thrombosis and Haemostasis<br>BLEEDING<br>criteria from International Society on Thrombosis and Haemostasis |  |
| Watt (2020) [20] | Retrospective cohort study | 1097 patients undergoing elective MVr for mitral regurgitation between April 2003 and March 2017<br>Excluded previous cardiac surgery and those with a history of atrial fibrillation, preoperative dialysis, bleeding or clotting disorder, or preoperative anticoagulation for any reason. Patients undergoing concomitant tricuspid valve repair (TVr) were included, whereas those undergoing concomitant coronary artery bypass grafting, aortic valve replacement, or left ventricular aneurysm repair were excluded. Patients who developed postoperative atrial fibrillation and remained in atrial fibrillation at the time of hospital discharge were also excluded from our analysis to reduce confounding in assessing anticoagulation among patients in normal sinus rhythm after MVr. | 1. Warfarin and aspirin 81mg for 30 days, then aspirin alone<br>2. Aspirin 81mg alone | FOLLOW UP<br>6 months<br>THROMBOEMBOLIC<br>pulmonary embolism, TIA, or stroke within 6 months after MVr<br>BLEEDING<br>postoperative pericardial effusion or cardiac tamponade, gastrointestinal (GI) bleeding, and reoperation for bleeding within 6 months after MVr.<br>OTHER<br>mortality and readmission within 6 months of MVr | Excluded in-hospital atrial fibrillation |

AF – atrial fibrillation; AHA – American Heart Association; ASA – aspirin (acetylsalicylic acid); BARC - Bleeding Academic Research Consortium; CVA – cerebrovascular accident; DOAC – direct oral anticoagulant; ISTH - International Society on Thrombosis and Haemostasis; MVr – mitral valve repair; TVr – tricuspid valve repair; TIA – transient ischaemic attack; VKA – vitamin K antagonist

**Table 2.**
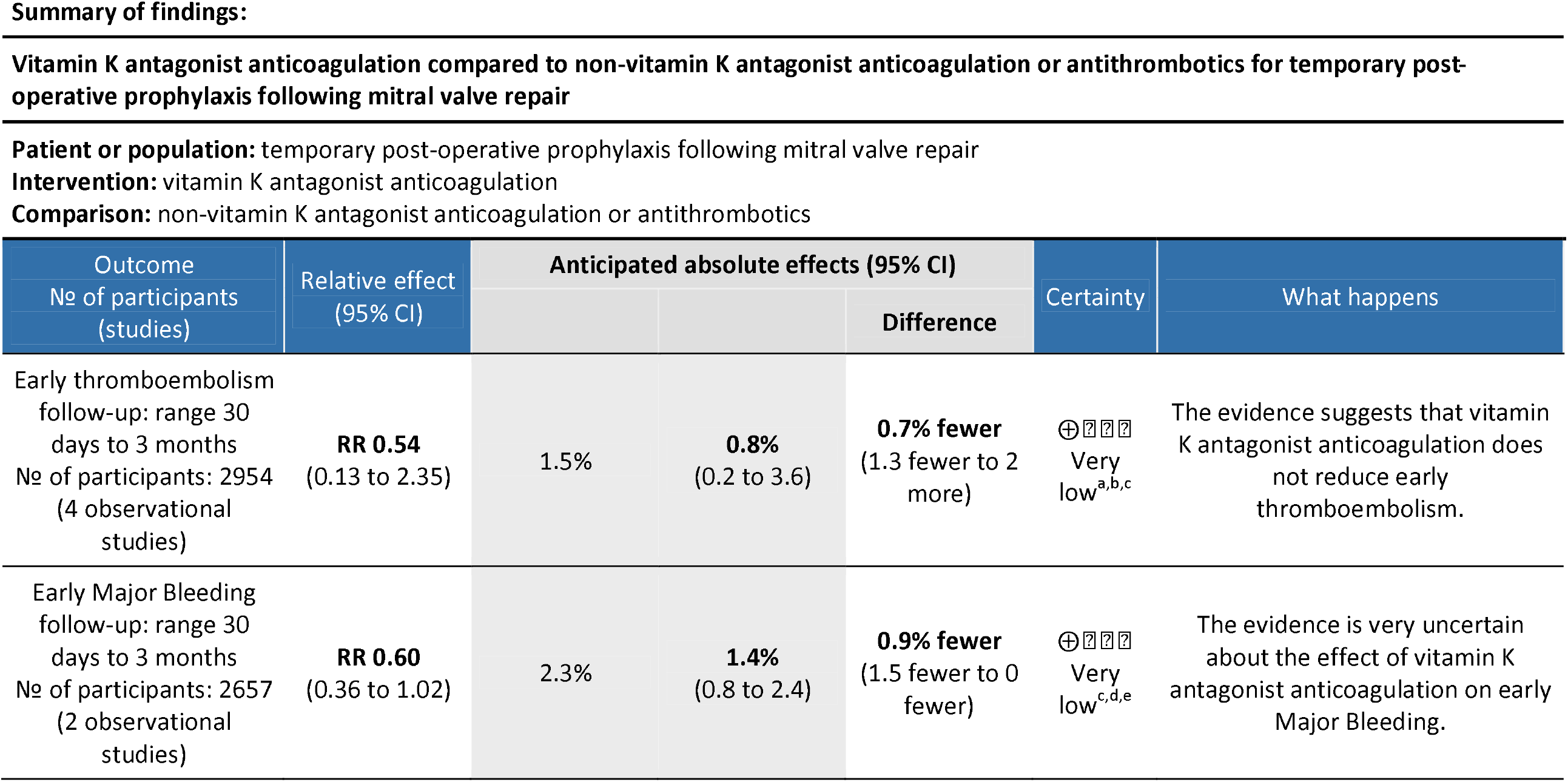
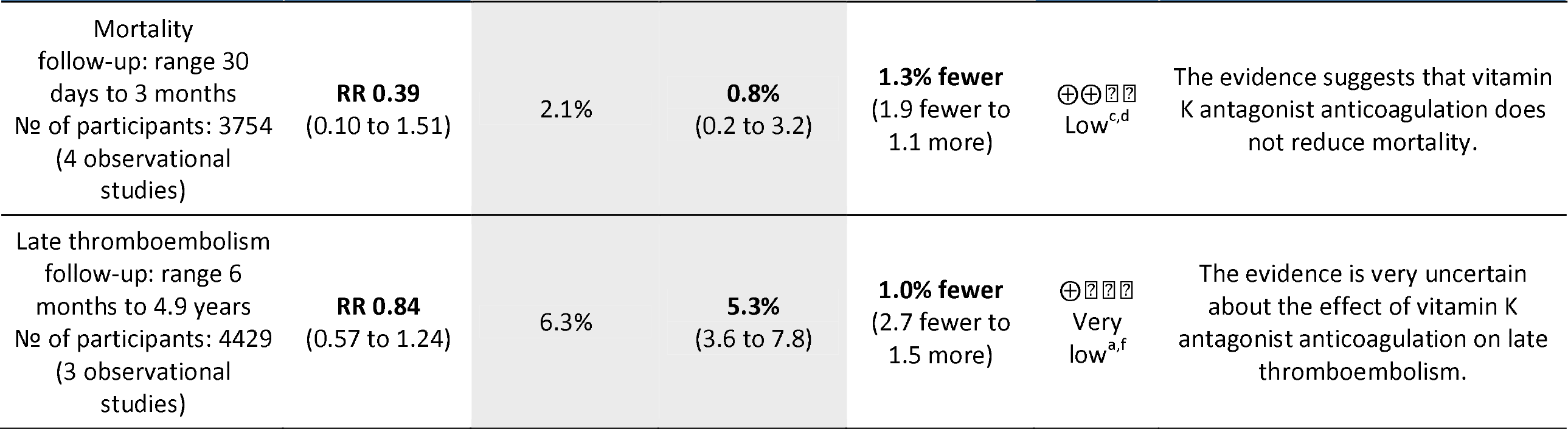
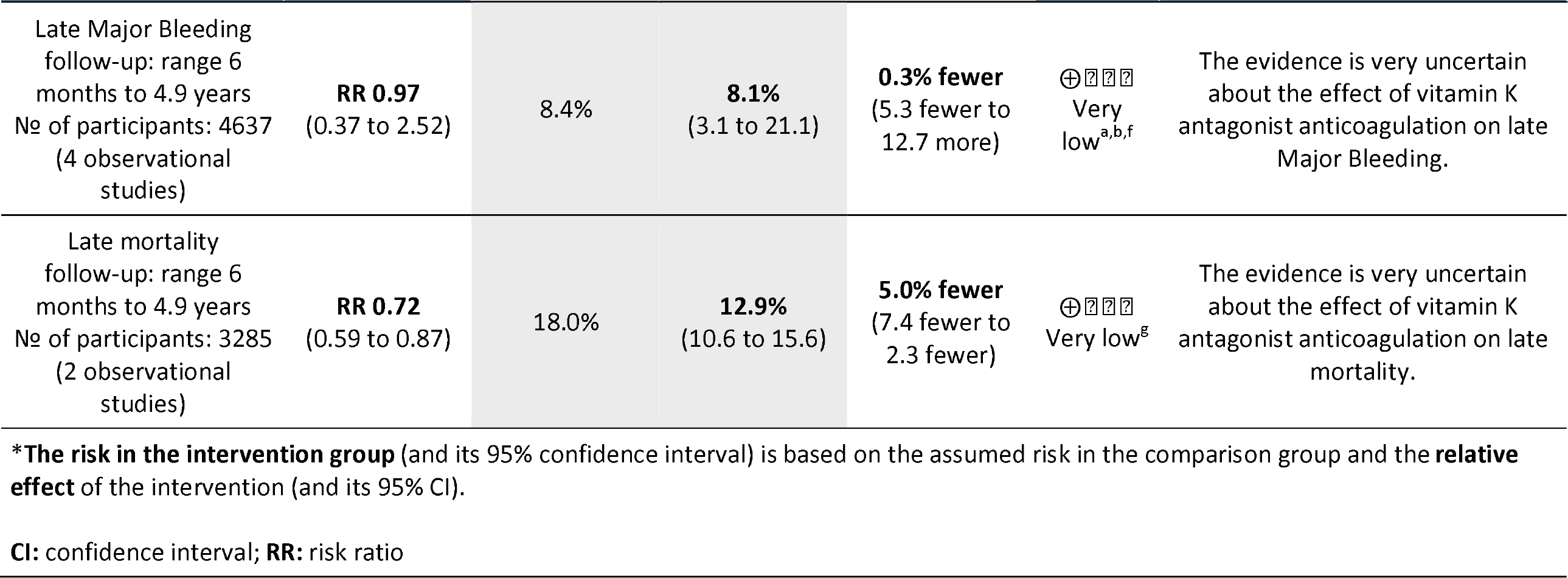
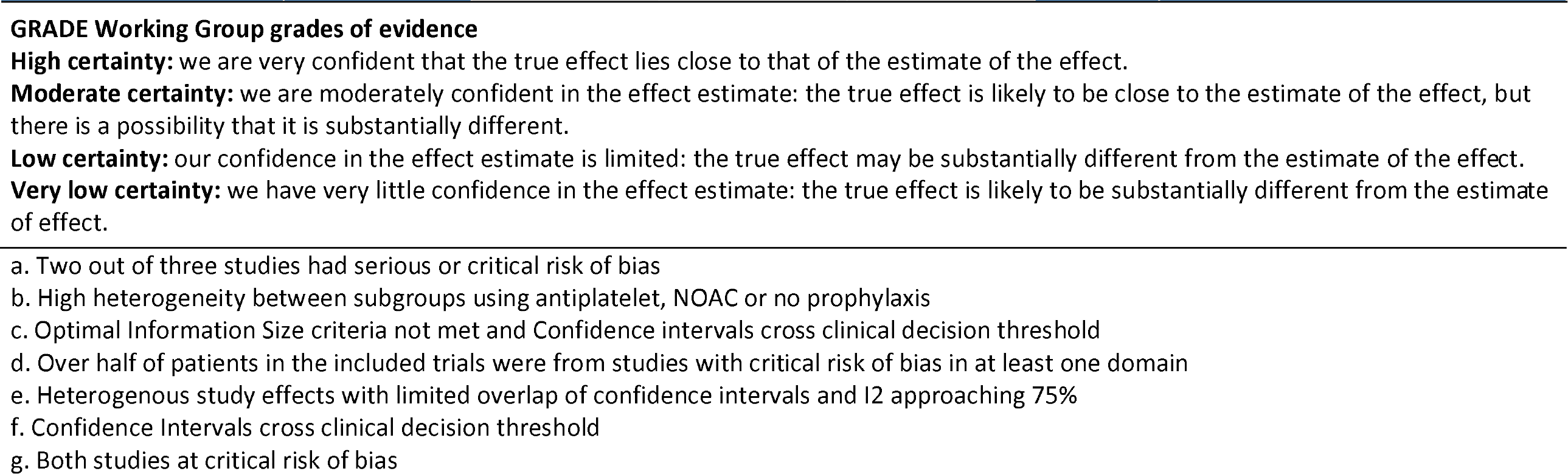
Summary of Findings Table

**Figure 1.**
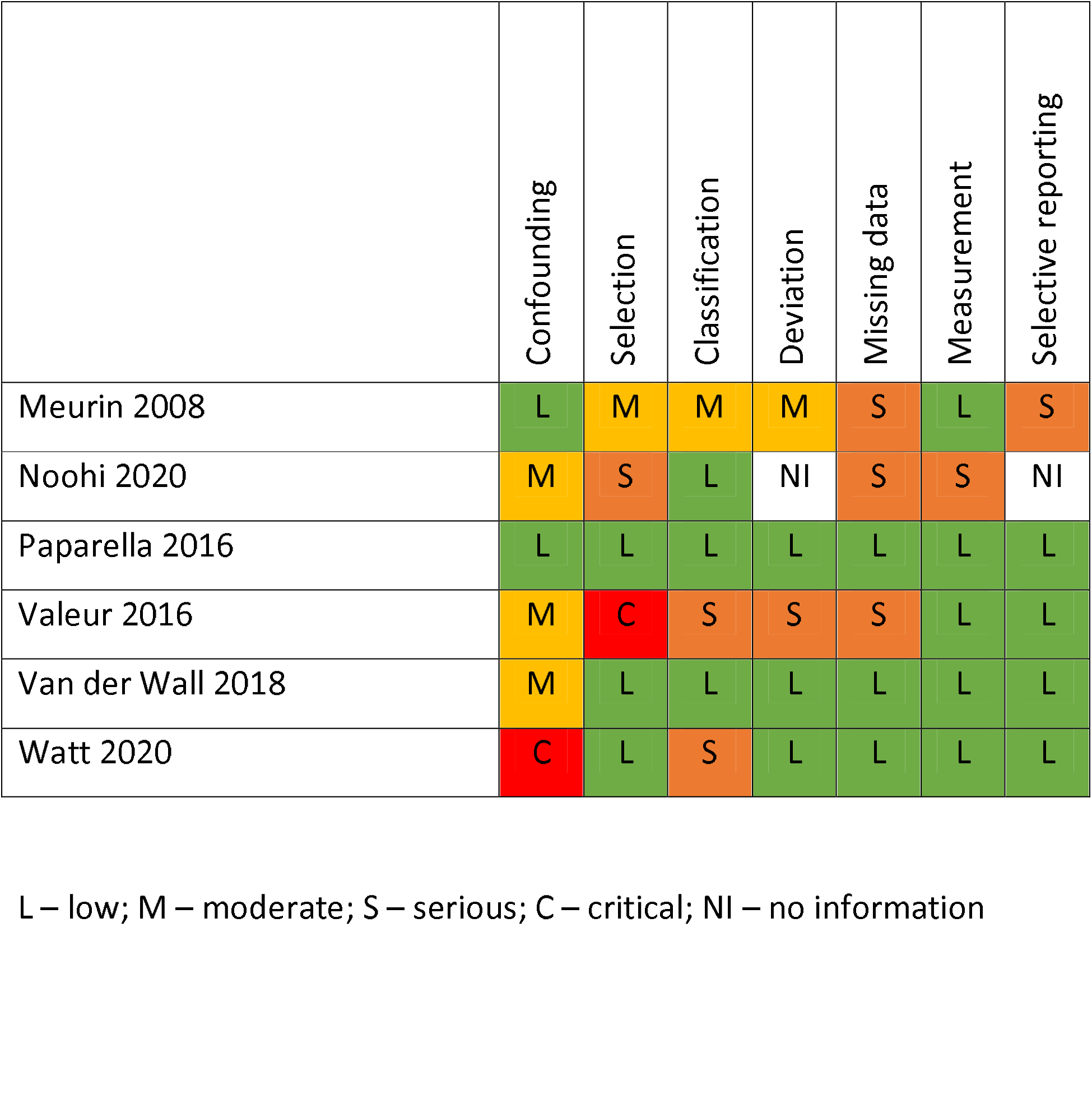
Risk of Bias summary

**Figure 2.**
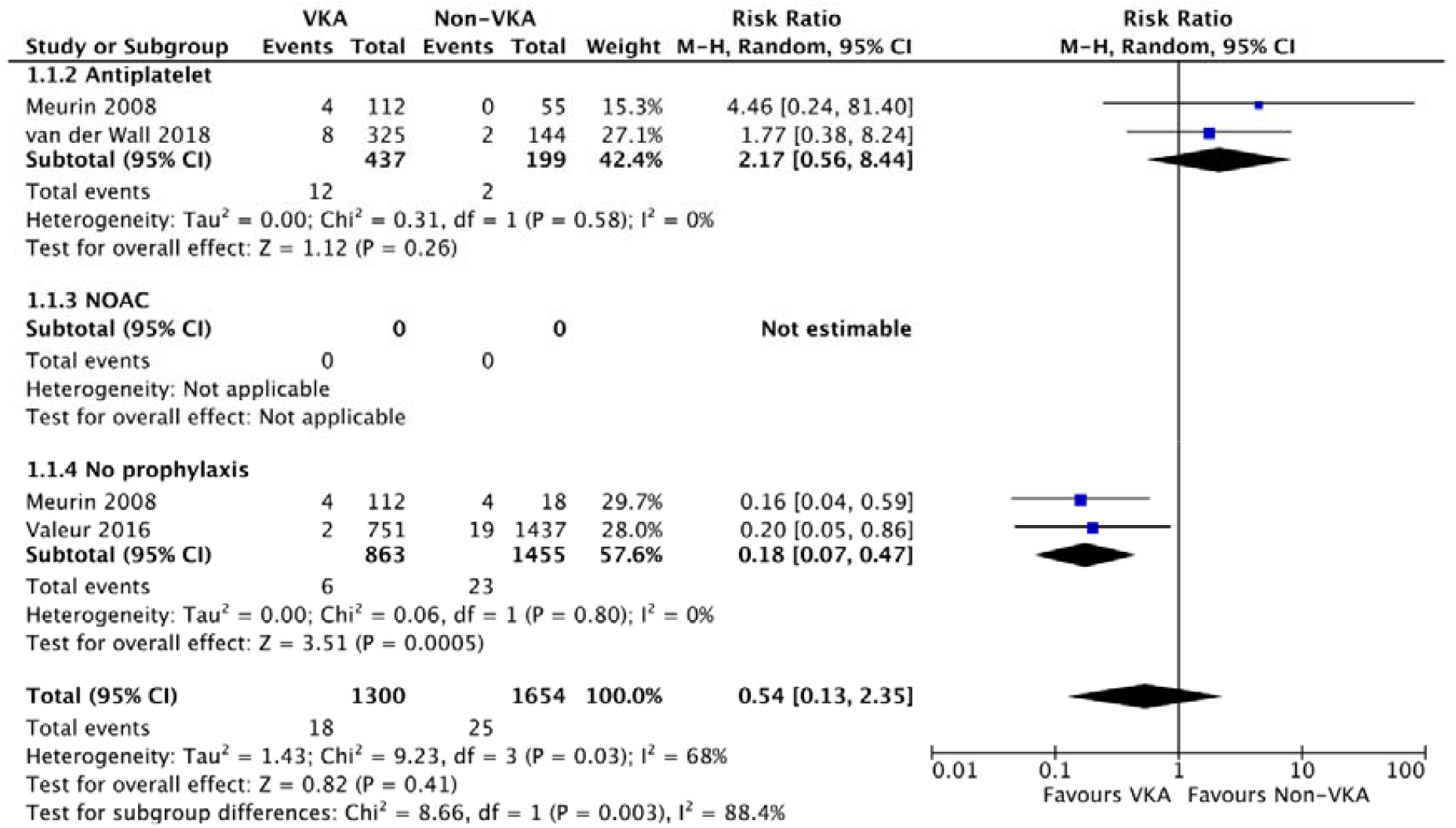
Forest plot of early thromboembolic risk comparing Vitamin K antagonist (VKA) anticoagulation with other strategies

**Figure 3.**
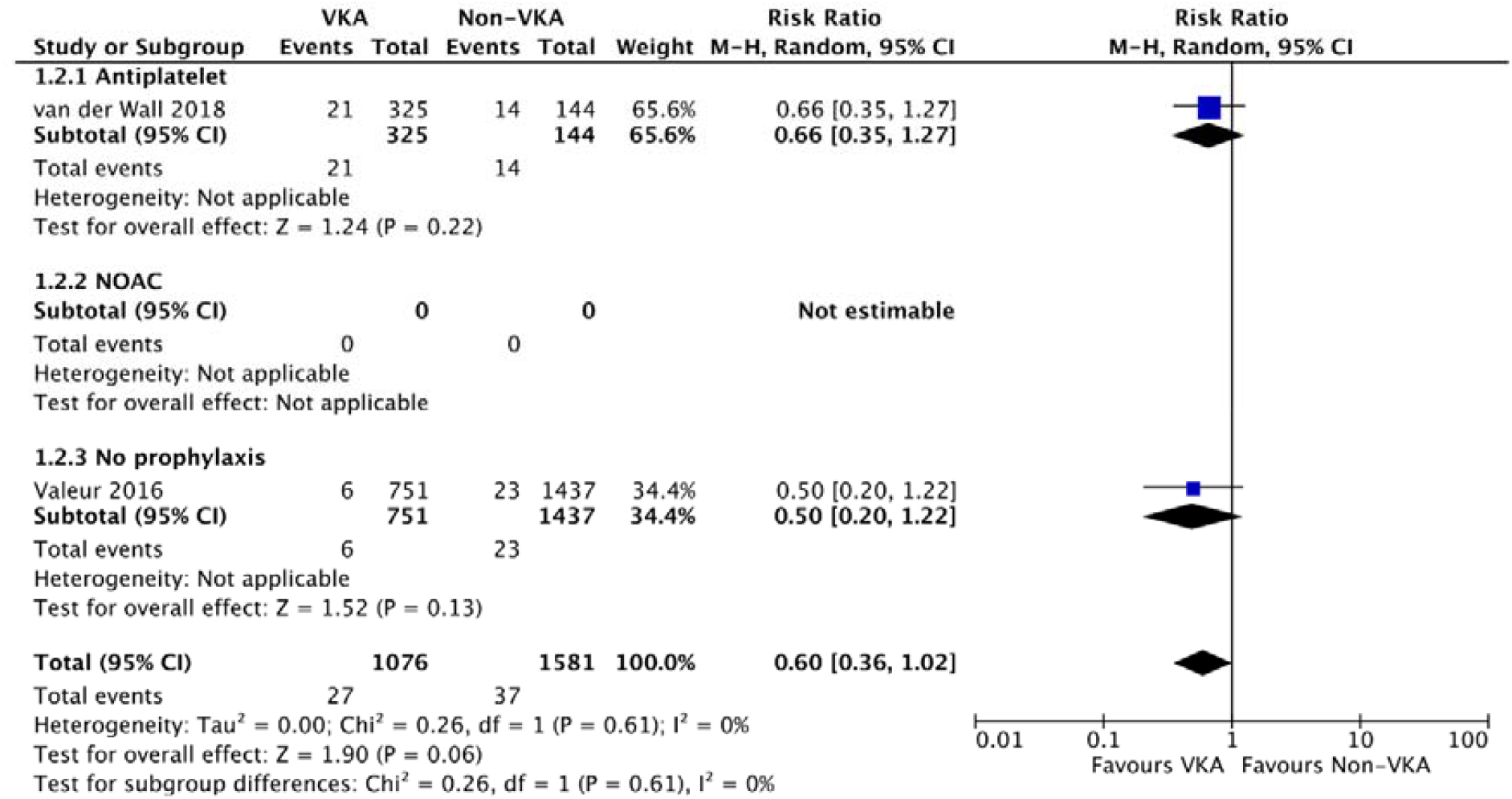
Forest plot of early major bleeding risk comparing Vitamin K antagonist (VKA) anticoagulation with other strategies

**Figure 4.**
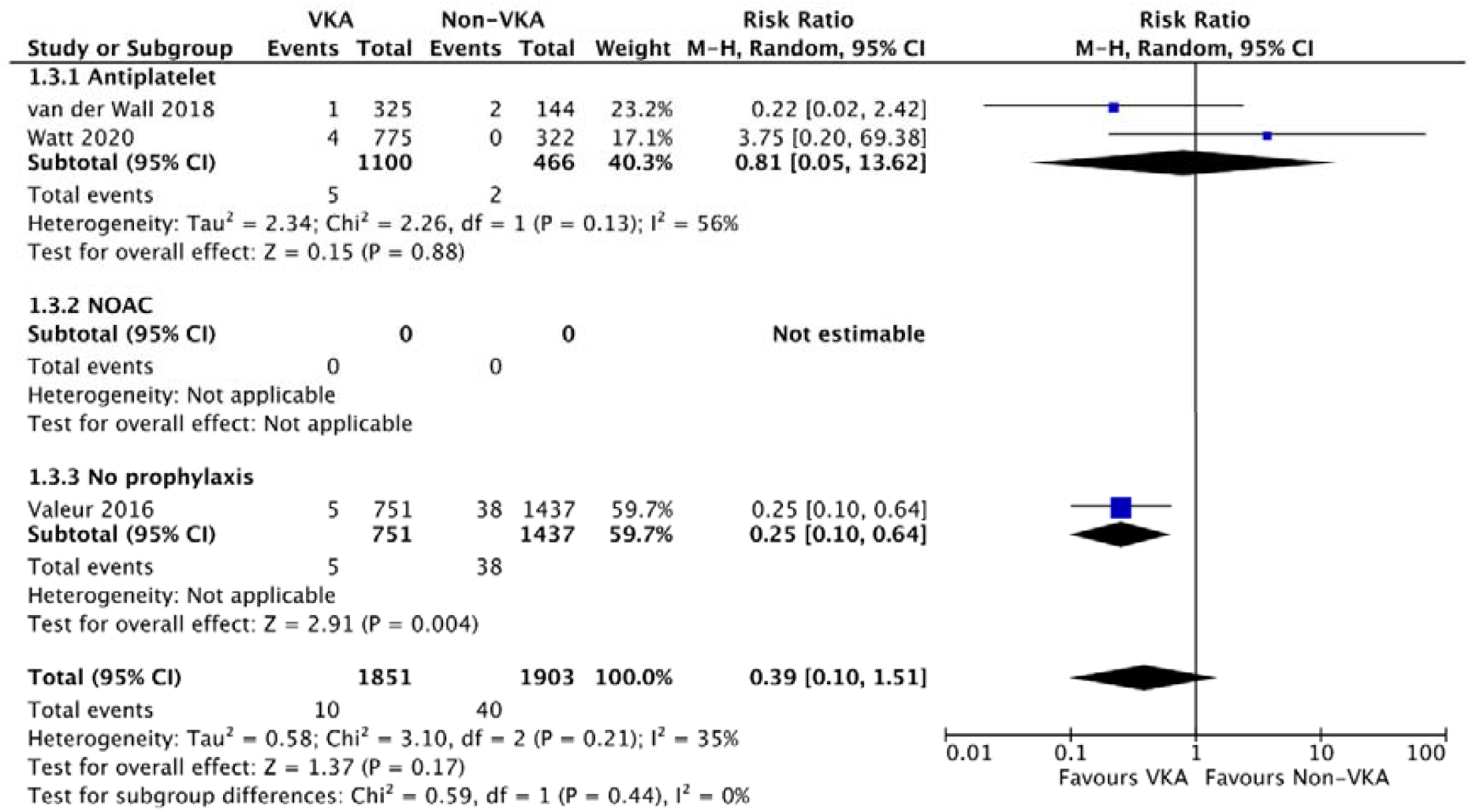
Forest plot of early mortality risk comparing Vitamin K antagonist (VKA) anticoagulation with other strategies

**Figure 5.**
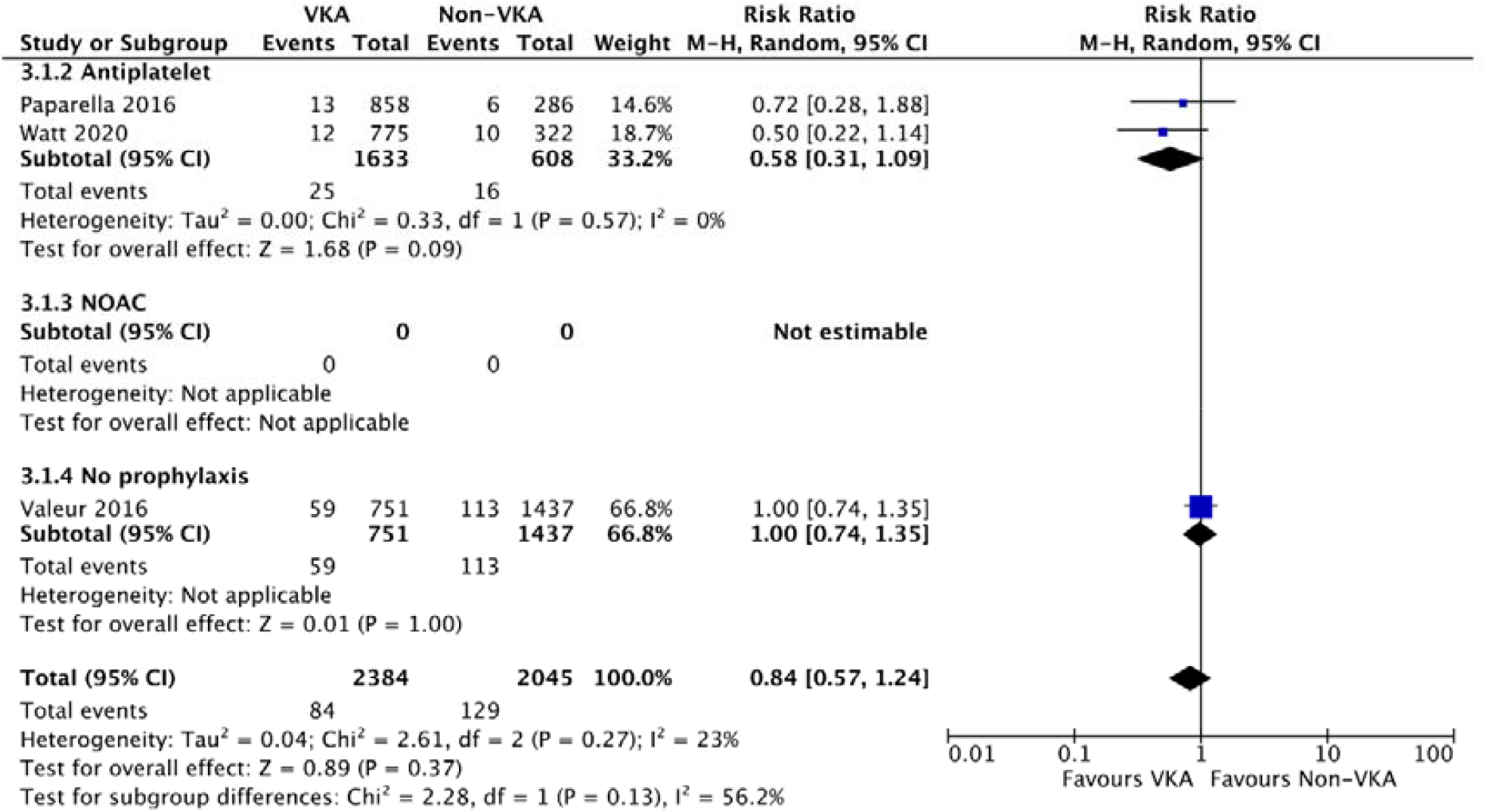
Forest plot of late thromboembolic risk comparing Vitamin K antagonist (VKA) anticoagulation with other strategies

**Figure 6.**
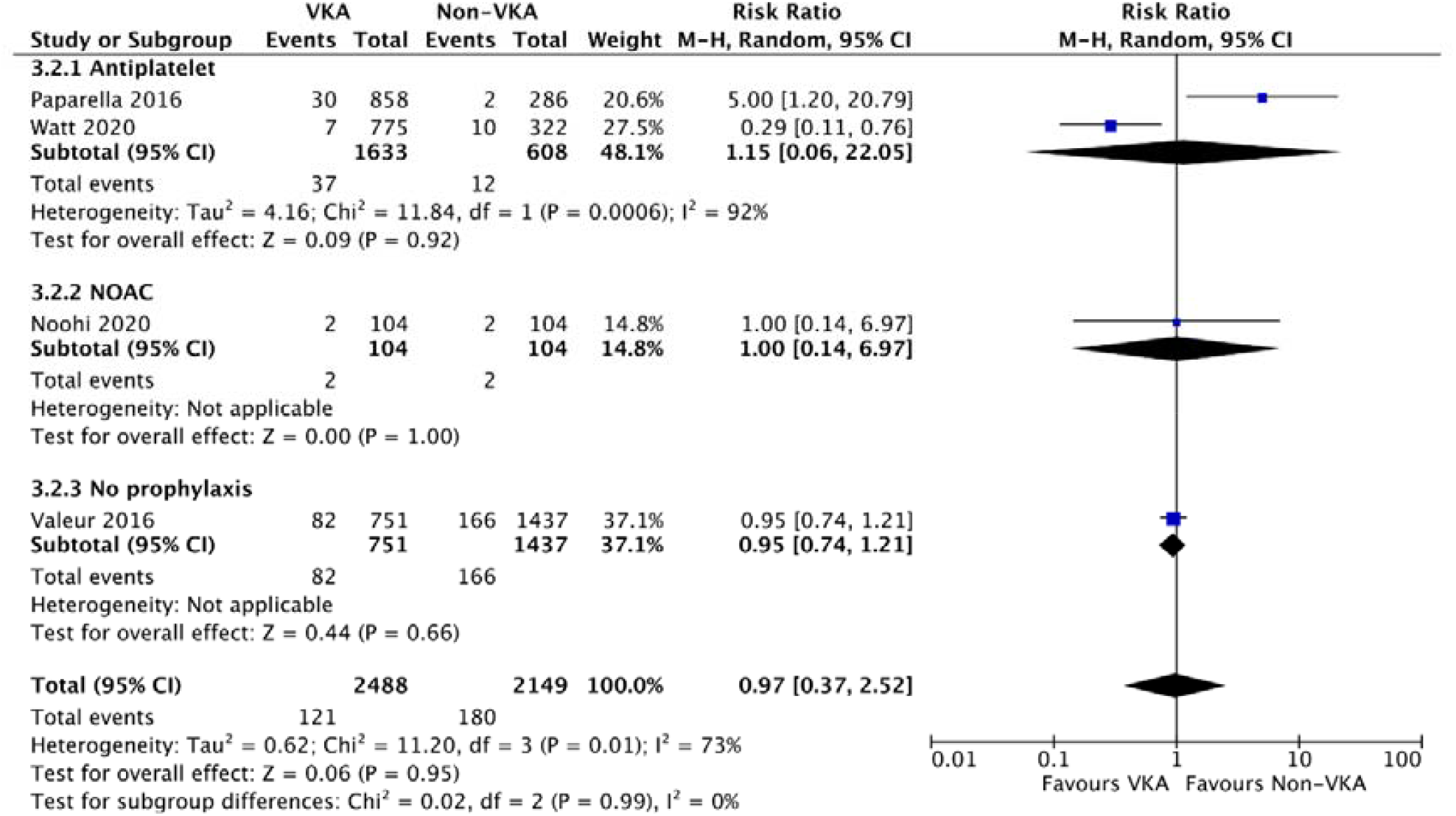
Forest plot of late major bleeding risk comparing Vitamin K antagonist (VKA) anticoagulation with other strategies

**Figure 7.**
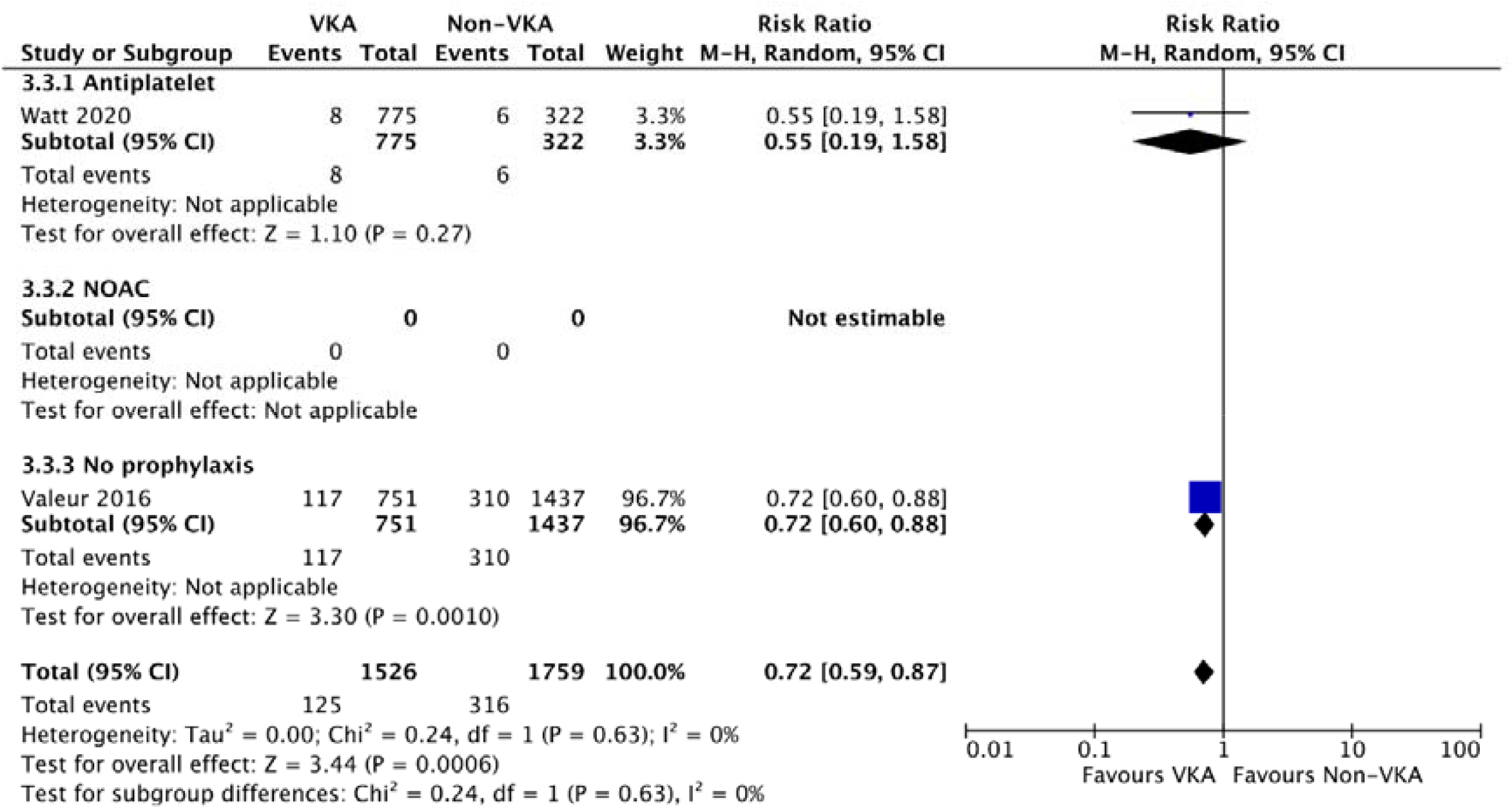
Forest plot of late mortality risk comparing Vitamin K antagonist (VKA) anticoagulation with other strategies

Due to the methodological and clinical diversity of the included studies, a random effects model was chosen for data synthesis. The Mantel-Haenszel method was employed as the event rate was low and study sizes generally small. This method has been demonstrated to have better statistical properties in such cases where data are sparse. Heterogeneity was assessed by visual inspection of the results as well as scrutiny of the I^2^ statistic for inconsistency, using an arbitrary cut-off of 75%. Statistical heterogeneity was assessed using subgroup analysis according to non-VKA strategy (i.e. DOAC, antiplatelet, or nil treatment). Given the small number of studies, further analysis of heterogeneity was assessed by examining individual study designs based on population characteristics such as relevant comorbidities, range of surgeries, and other plausible causes for variation. These were described in narrative format in the discussion. Sensitivity analyses were performed excluding studies with any critical risk of bias in any domain or where the cumulative effect of multiple domains graded as serious risk of bias was considered sufficient to grade the study as a whole at critical risk of bias.

### Reporting bias assessment

We intended to undertake funnel plot analysis where there were more than 10 studies included in the data synthesis. For any prospective randomized trials, we intended to compare trial protocols with the published outcomes to assess for any reporting bias. Due to study paucity, funnel plot analyses were not required. Likewise, because of the absence of prospective randomised trials, protocols did not need to be reviewed.

### Certainty assessment

Two reviewers (BHK and SGJ) assessed the certainty of the evidence against GRADE criteria, using the Cochrane Handbook recommendations for methodology and GRADEPro software for constructing a Summary of Findings table.

## Results

A PRISMA Flow Diagram demonstrates the search and selection process (Figure 8). There were no randomised controlled trials that fulfilled the inclusion criteria and six observational studies that were ultimately included in the data synthesis (15–20). Studies that were excluded following screening are listed (with reasons) in Appendix II. Included studies and their key characteristics are shown in Table 1. The risk of bias was assessed using the seven domains of the ROBINS-I tool and presented for each individual study following description of the study characteristics (Table 1) and again in summary format (Figure 1).

**Figure 8.**
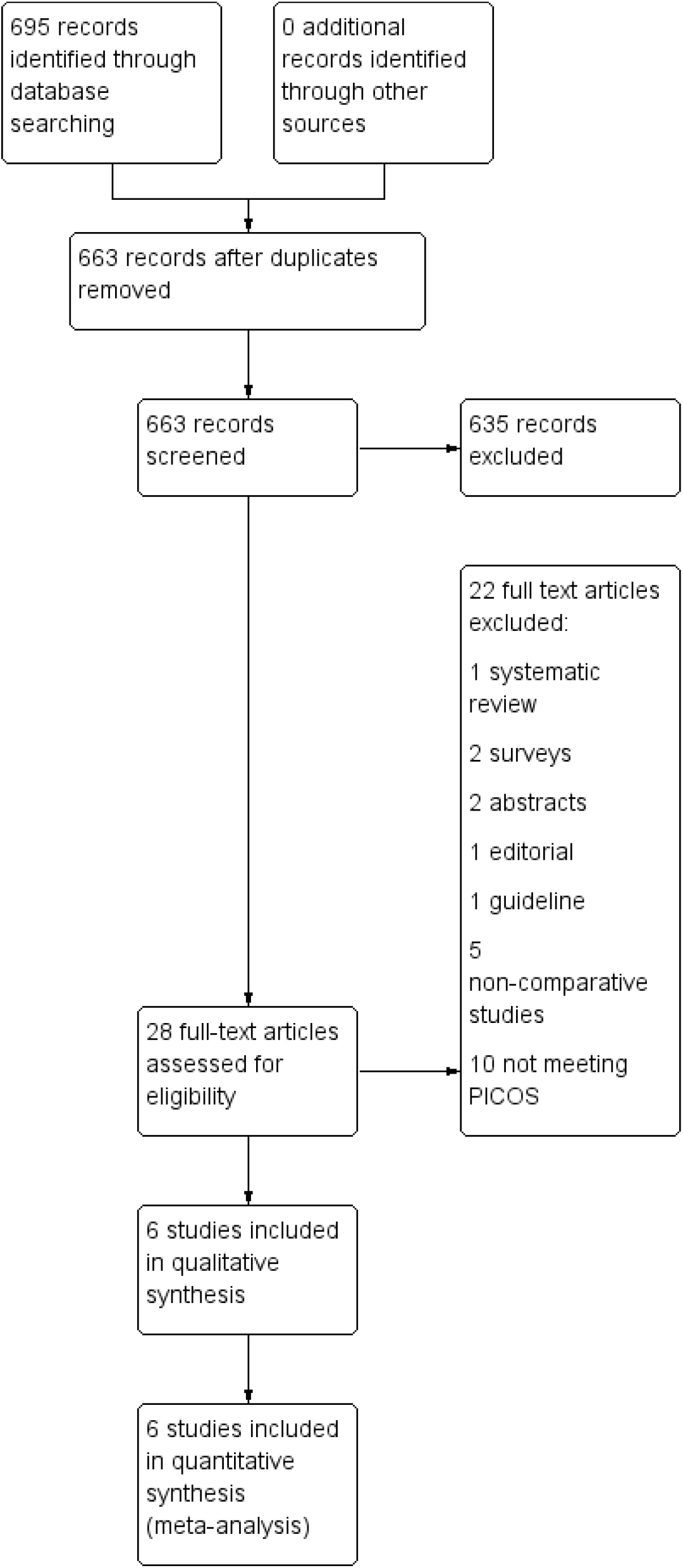
PRISMA Flow Diagram

All data were acquired from primary sources in unmanipulated format unless otherwise specified in the study descriptions. The summary statistics for each study are included in the forest plots for each outcome, showing effect estimate and 95% confidence interval.

### Thromboembolism

Six observational cohort studies (15-20) reported thromboembolic complications following mitral valve repair, comparing the effects of VKA anticoagulation with non-VKA anticoagulation, antiplatelet treatment or no treatment. Three studies (15,18-19) reported on early thromboembolic risk and three studies (17-18,20) on late risk (mean followup 6-12m). A total of 5291 patients were recruited with 2925 in the VKA treatment arm, 807 in the antiplatelet arm, 104 in the DOAC arm and 1455 in the no treatment arm. One study (15) had three arms and was therefore included in two subgroups against VKA, reflected in the higher patient numbers in the summary of findings table as the VKA arm patients were counted in two subgroups. Two studies (17,19) comparing VKA to antiplatelet treatment had low risk of bias, one (15) had serious risks of bias and one (20) had critical risk of bias in the confounding domain. There were no events in either arm in the only VKA vs DOAC study (16) and zero cell corrections made no difference to the effect estimates. Within the VKA vs no treatment subgroup, one study had critical risk of bias in the selection domain due to exclusion of patients suffering stroke within seven days of discharge (18). When this was excluded in sensitivity analysis, one study which did not meet Optimal Information Size criteria (15) was left in the subgroup and the effect estimate based on this one study was in favour of VKA treatment based on thromboembolic complications.

Overall, the impact of VKA use (as compared to antiplatelet therapy or no treatment) on the risk of thromboembolic complications, predominantly ischemic stroke, was uncertain given that the quality of the evidence was very low and the effect estimate did not exclude both significant benefit or harm. This was the case for both early (relative risk [RR] 0.54, 95% CI 0.13 – 2.35, p=0.41, I^2^ = 68%, absolute risk difference 7 fewer per 1,000 (from 13 fewer to 20 more), overall certainty of evidence = very low) and late risk (RR 0.84, 95% CI 0.57 – 1.24, p=0.37, I^2^ = 23%, absolute risk difference 10 fewer per 1,000 (from from 27 fewer to 15 more, certainty of evidence = very low). Sensitivity analysis, excluding the studies with critical risk of bias did not change the effect estimates for either early or late thromboembolism.

### Major bleeding

Five studies (16-20) with a total of 5106 patients reported on major bleeding complications following mitral valve repair, comparing post-operative treatment in 2813 patients with VKA anticoagulation against 752 with antiplatelets, 104 with DOAC and 1437 with no treatment. Two studies (19-20) reported on early major bleeding and four (16-18,20) on late bleeding. Two studies in the antiplatelet subgroup were at low risk of bias and one (20) had one domain at critical risk of bias. The single study comparing DOACs against VKA had no events in either arm and was therefore excluded from effect estimation, except for sensitivity using zero-cell adjustments. Only one study comparing VKA with no anticoagulant or antithrombotic treatment reported major bleeding outcomes (18) and although this did not show any difference between the two strategies, this was at critical risk of bias in the selection domain for excluding patients with events within 7 days of discharge.

VKA use may be associated with no difference in the risk of early major bleeding complications, with a wide confidence interval spanning both risk and harm, although the evidence is very uncertain (RR 0.60, 95% CI 0.36 – 1.02, p=0.06, I^2^=0%, absolute risk difference 9 fewer per 1,000 (from 15 fewer to 0 fewer), certainty of evidence = very low). Similarly, the effect of VKA use on late major bleeding events was also uncertain (RR 0.97, 95% CI 0.37 – 2.52, p=0.95, I^2^=73%, absolute risk difference 3 fewer per 1,000 (from from 53 fewer to 127 more, certainty of evidence = very low). Sensitivity analysis, removing the studies at critical risk of bias, did not change the overall effect estimates.

### Mortality

Four studies (17-20) reported mortality following mitral valve repair, including 4898 patients. Of these, 2709 received VKA anticoagulation, 752 received antiplatelets and 1437 received no treatment. Three studies (18-20) reported early mortality and three studies (17-18,20) reported late mortality. No studies reporting this outcome included NOACs in the intervention arm. In the antiplatelet arm, two studies (17,19) were at low risk of bias and one (20) had critical risk of bias in the confounding domain for strong associations between operating surgeon, operative time, and anticoagulant use. Only one study compared VKA anticoagulation against no treatment (18), favouring VKA, but this was at critical risk of selection bias from exclusion of patients with events prior to seven days after discharge.

VKA use had an unclear effect on early mortality, as the confidence intervals span risk and benefit (RR 0.39, 95% CI 0.10 – 1.51, p=0.17, I^2^=35%, absolute risk difference 13 fewer per 1,000 (from 19 fewer to 1 more), certainty of evidence = low). Sensitivity analysis did not change the overall effect. For late mortality, there may have been a small risk reduction with VKA anticoagulation, although the evidence was very uncertain (RR 0.72, 95% CI 0.59 – 0.87, p=0.0006, I^2^=0%, absolute risk difference 50 fewer per 1,000 (from 74 fewer to 2 fewer, certainty of evidence = very low). Sensitivity analysis, removing the study at critical risk of bias from this analysis left only one study, which did not show any advantage of VKA anticoagulation in the long term.

## Discussion

The results of this systematic review and meta-analysis confirm that there is insufficient evidence to judge the optimal treatment strategy to minimize thromboembolic complications following mitral valve repair. There was also insufficient evidence of good quality to make a judgement on the relative risks of VKA anticoagulation versus NOACs or versus no treatment. One study (17) comparing VKA anticoagulation against antiplatelet treatment was sufficiently well designed as to be considered at low risk of bias in all domains, including confounding which is ordinarily moderate risk of bias in the ROBINS-I tool for observational studies. Another was low risk in all but confounding (19). Nonetheless, they were still non-randomized and had low numbers of events and were not supported by sufficient other studies of good quality to give confidence in the certainty of the evidence. The small effect of VKA on late mortality may be due to be the result of residual confounders from the study designs.

Overall, this review confirms an ongoing need to demonstrate the effectiveness and long-term safety of VKA anticoagulation following mitral valve repair as many of the effect estimates cross the clinical decision threshold and all domains had low or very low certainty of evidence. The heterogenous practices in anticoagulation and antithrombotic management demonstrated in international surveys indicates clinician uncertainty in the validity of guidelines. Furthermore, the trend of anticoagulation with NOACs seen in more recent (pending publication) surveys, confirms that further studies are required to establish the appropriate management strategy for these patients.

Most studies appropriately excluded concomitant procedures that might have confounded the results. It was unclear in most studies whether the analysis was based on the initial treatment regimen planned, or the one at final follow-up. As the rate of atrial fibrillation in post-cardiac surgery patients approaches 30%, it is likely that a proportion of patients in retrospective studies initially intended for antiplatelet management alone were subsequently prescribed anticoagulants. Some patients were also prescribed VKA anticoagulants in addition to aspirin, and this was variably recorded. The exclusion of patients in atrial fibrillation at the time of discharge from hospital or the inclusion of these patients in the VKA anticoagulation arm of the studies may have further skewed the results. Furthermore, the decision not to use VKA anticoagulation in observational studies may have been clinically guided (e.g. in patients with bleeding diathesis), leading to selection bias. The higher bleeding rates in the non-VKA group of the study by Valeur et al may be an indication of this type of clinical decision.

Whilst the patient populations between studies were similar, event rates between studies were low and heterogenous. No studies included risk stratification for thrombosis or bleeding such as CHA_2_DS_2_-VASc, HAS-BLED or ORBIT scoring. Heterogeneity is also likely to have been affected by differing time points of follow-up for various studies, variations in precise definitions of outcome measures (e.g. the use of ischaemic stroke as a surrogate for systemic thromboembolism in several studies will have excluded non-cerebral organ embolization; and the inclusion of pulmonary thromboembolism in one study, which is not normally associated with left-sided heart valve thrombosis, will also have affected the comparisons).

This review was written to inform the design of an international prospective multi-centre randomized controlled trial. This would be designed as a pragmatic study, accounting for confounders, including the propensity of mitral valve repair patients to atrial fibrillation, and the three treatment modalities. To answer the study question effectively, this would need to be powered to identify events in each arm accounting for the invariable cross-over between arms, mitigating for the risks of bias seen in these observational studies.

### Limitations

It is a significant limitation of this study that VKA anticoagulation was compared against NOAC, aspirin and no-treatment. Both within individual studies and between studies for this meta-analysis, these three treatments were grouped whereas they are sufficiently different in action and effect to be incomparable with each other. Some mitigation was provided in subgrouping the effects of these heterogenous treatment arms, but the aggregation of effect is a significant design limitation, performed here only due to the lack of studies.

In conclusion, there is insufficient evidence of good quality to support the current guidelines recommending use of VKA anticoagulation in patients following mitral valve repair, and more robust evidence is required to establish the optimal antithrombotic regime.

## Data Availability

All data produced in the present work are contained in the manuscript

## Appendix I Search Strategies

MEDLINE via PubMed

“mitral AND (repair OR ring OR annuloplasty) AND (anticoag* OR antiplat* OR aspirin OR clopidogrel OR prasugrel OR ticagrelor OR DAPT OR SAPT OR thienopyridine OR warfarin OR coumarin OR VKA OR “vitamin K antagonist” OR NOAC OR “novel oral anticoag*” OR DOAC OR “direct oral anticoag*” OR edoxaban OR apixaban OR rivaroxaban OR dabigatran)”

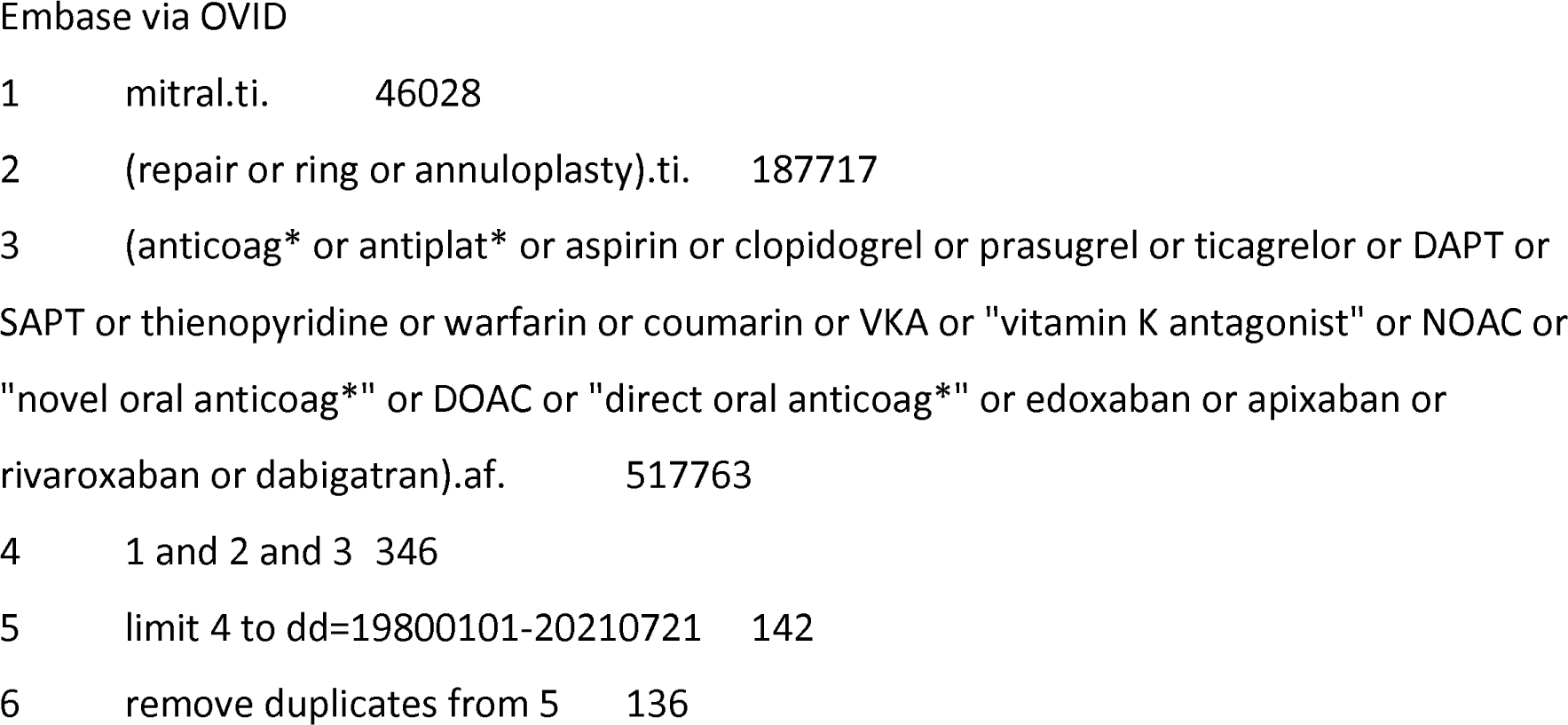

## Appendix II Excluded studies

Appendix II – Excluded studies
| Study | Reason for exclusion |
| --- | --- |
| Acanfora 2016 | Case series with no control group and non-repair cases |
| Aramendi 1998 | Mixed repairs and replacement comparing ticlopidine, aspirin, coumadin and no treatment |
| Bisson 2018 | All patients with pre-op AF (i.e. other indication for anticoagulation) |
| Chua 1994 | Case series with no control group and pre-op AF |
| Deloche 1990 | Case series with no control group and unclear warfarin strategy |
| Durães 2016 | Bioprosthetic mitral or aortic valves only |
| Espinoza 2018 | Abstract only |
| Galloway 1988 | Case series with no control group and concomitant surgery |
| Handa 1999 | All patients with pre-op AF (i.e. other indication for anticoagulation) |
| Jovin 2005 | All patients with post-op AF (i.e. other indication for anticoagulation) |
| Kim 2014 | All patients with pre-op AF (i.e. other indication for anticoagulation) |
| Papadimas 2021 | Systematic review |
| Potpara 2016 | Survey of practice |
| Russo 2018 | All patients with pre-op AF (i.e. other indication for anticoagulation) |
| Sadeghipour 2020 | Abstract only |
| Sternik 2016 | Opinion piece |
| Suri 2013 | Predictors of warfarin use (i.e. not comparative study) |
| Thomas 2017 | All patients with pre-op AF (i.e. other indication for anticoagulation) |
| Thourani 2013 | Combined repairs and replacements and concomitant surgery |
| Van der Wall 2018a | Erratum |
| Vaughan 2005 | Survey of practice |
| Whitlock 2005 | Guideline |

## Appendix III Details of Included Studies Meurin 2008

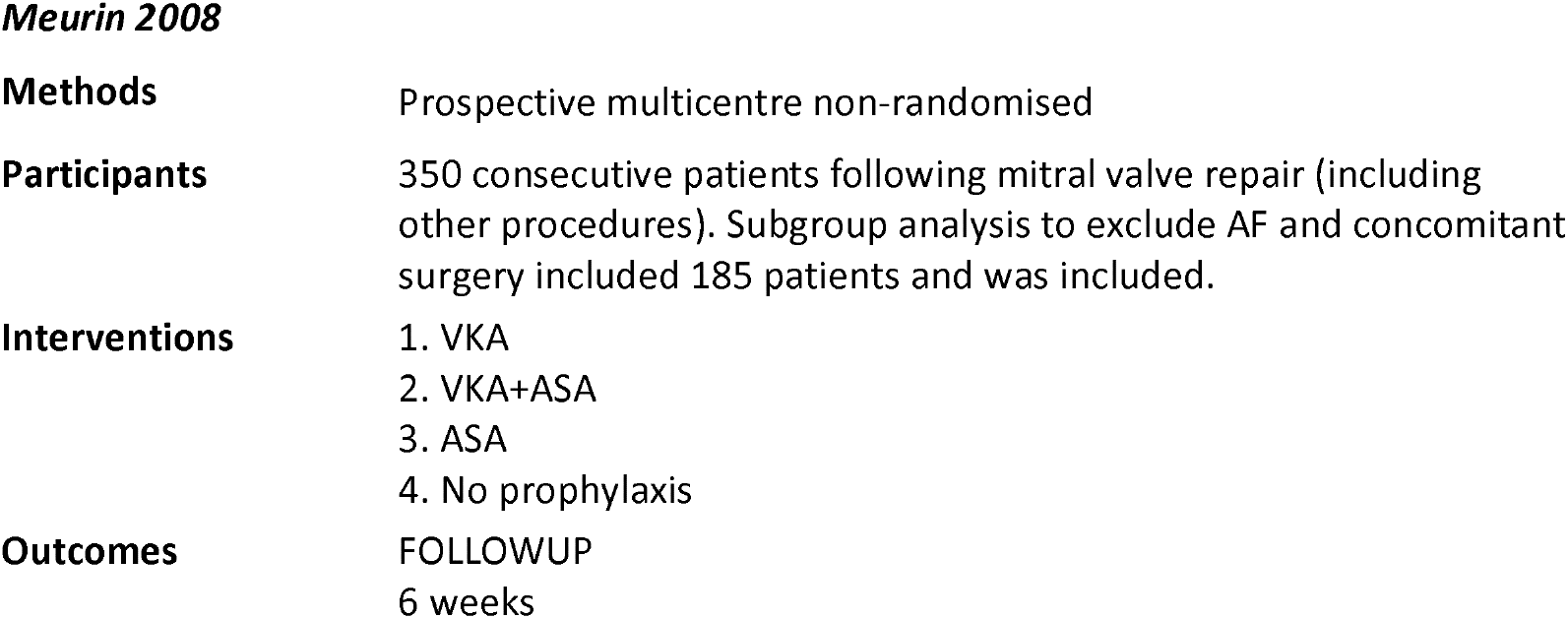
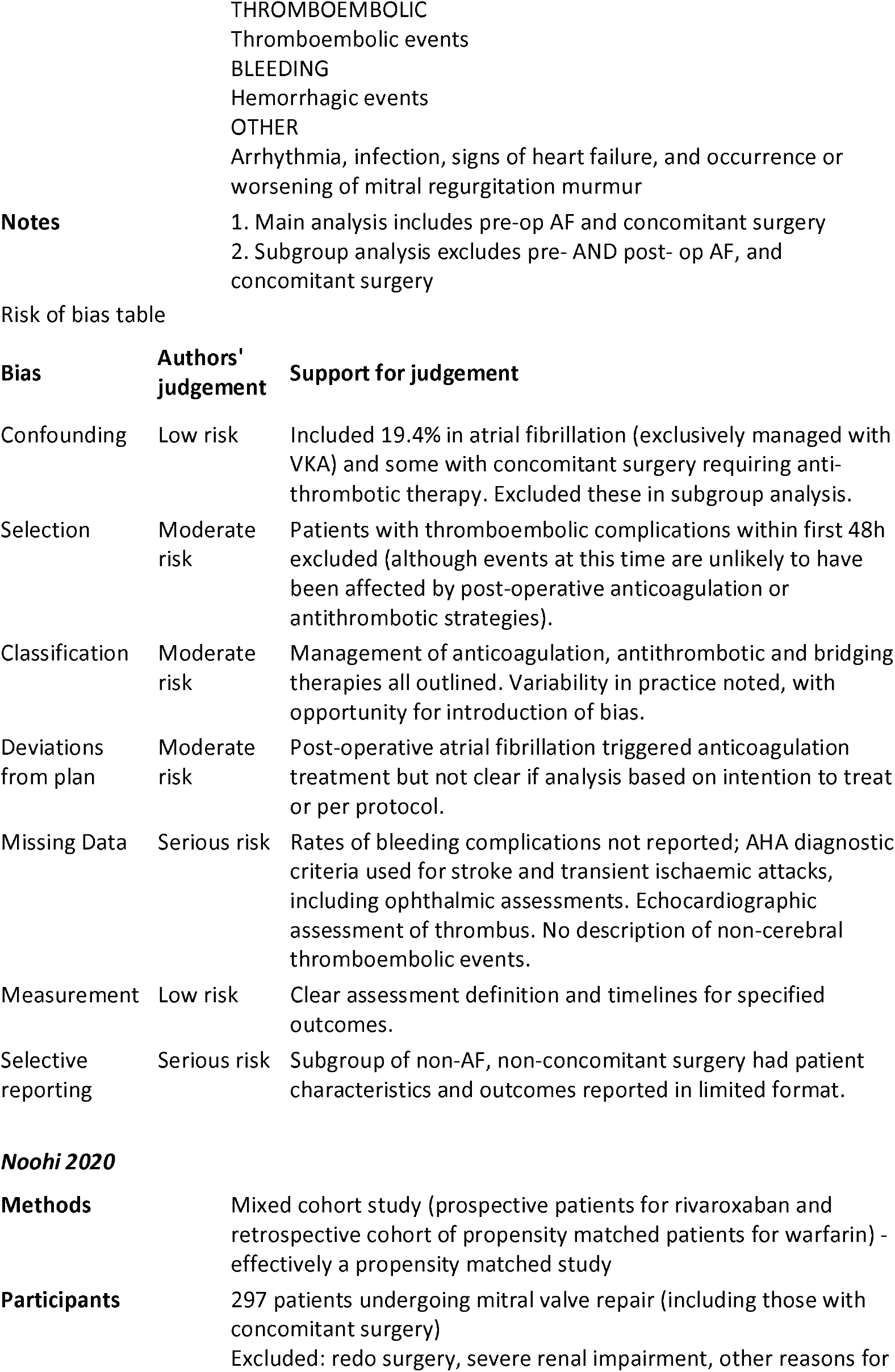
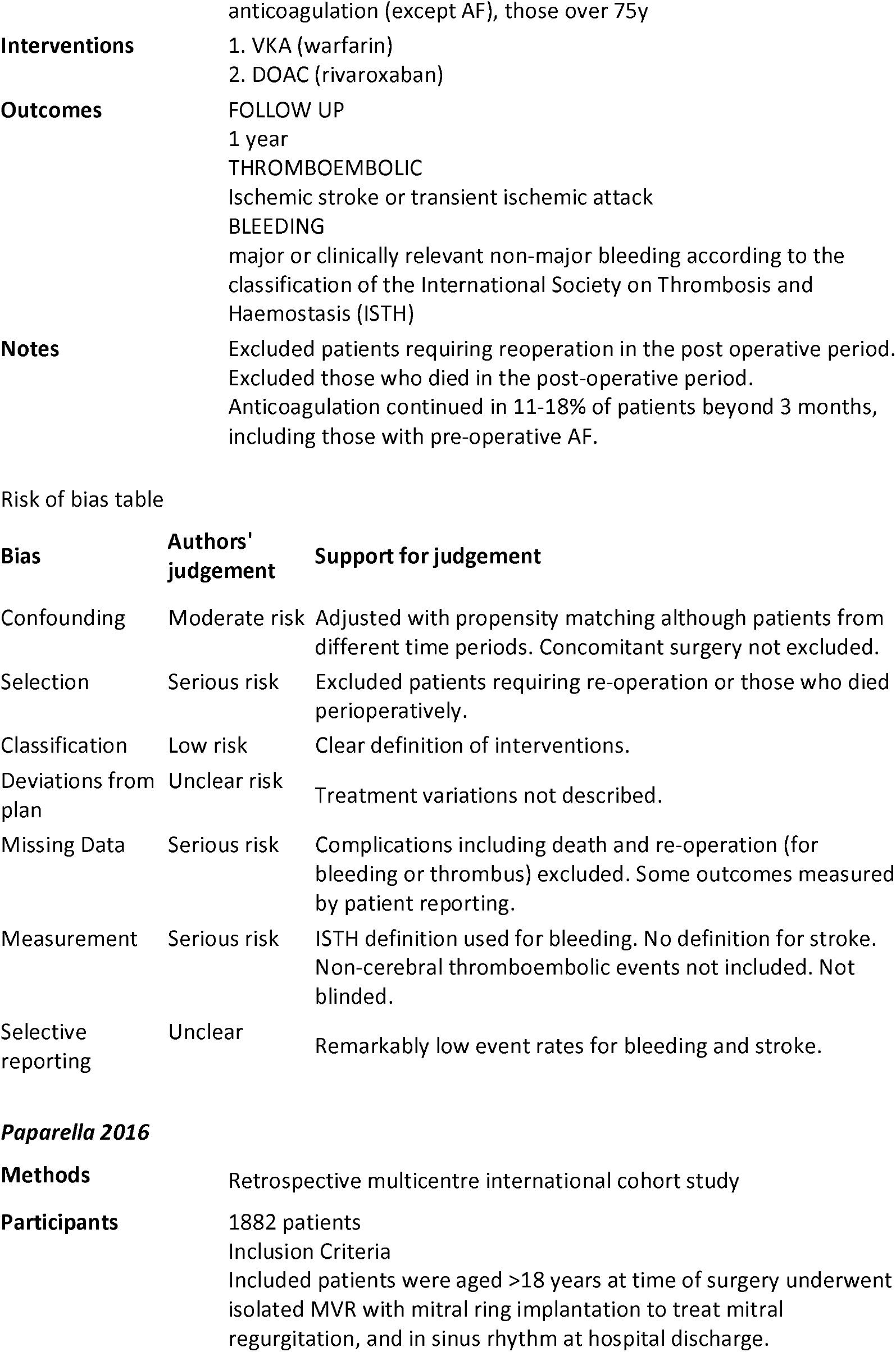
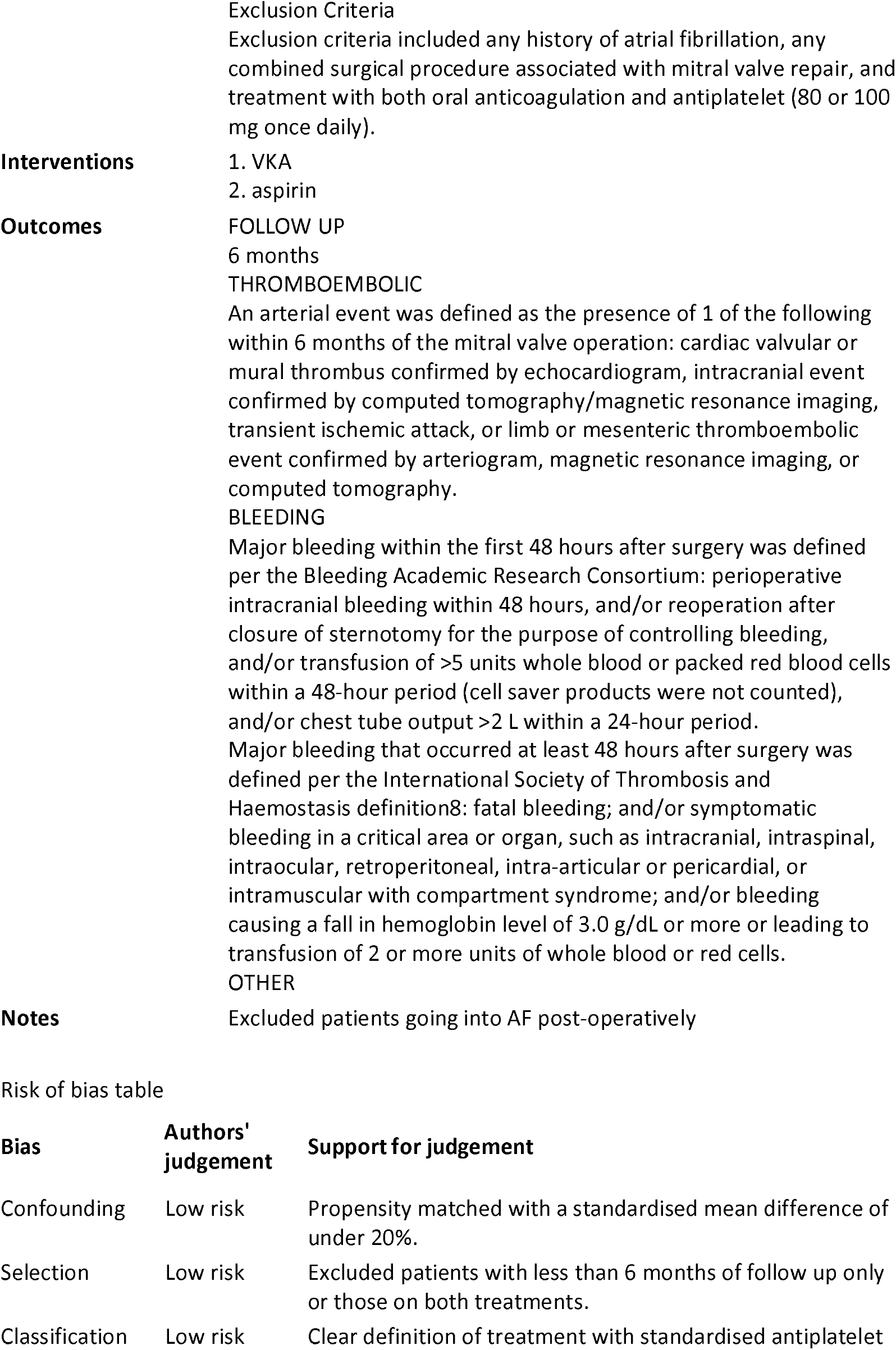
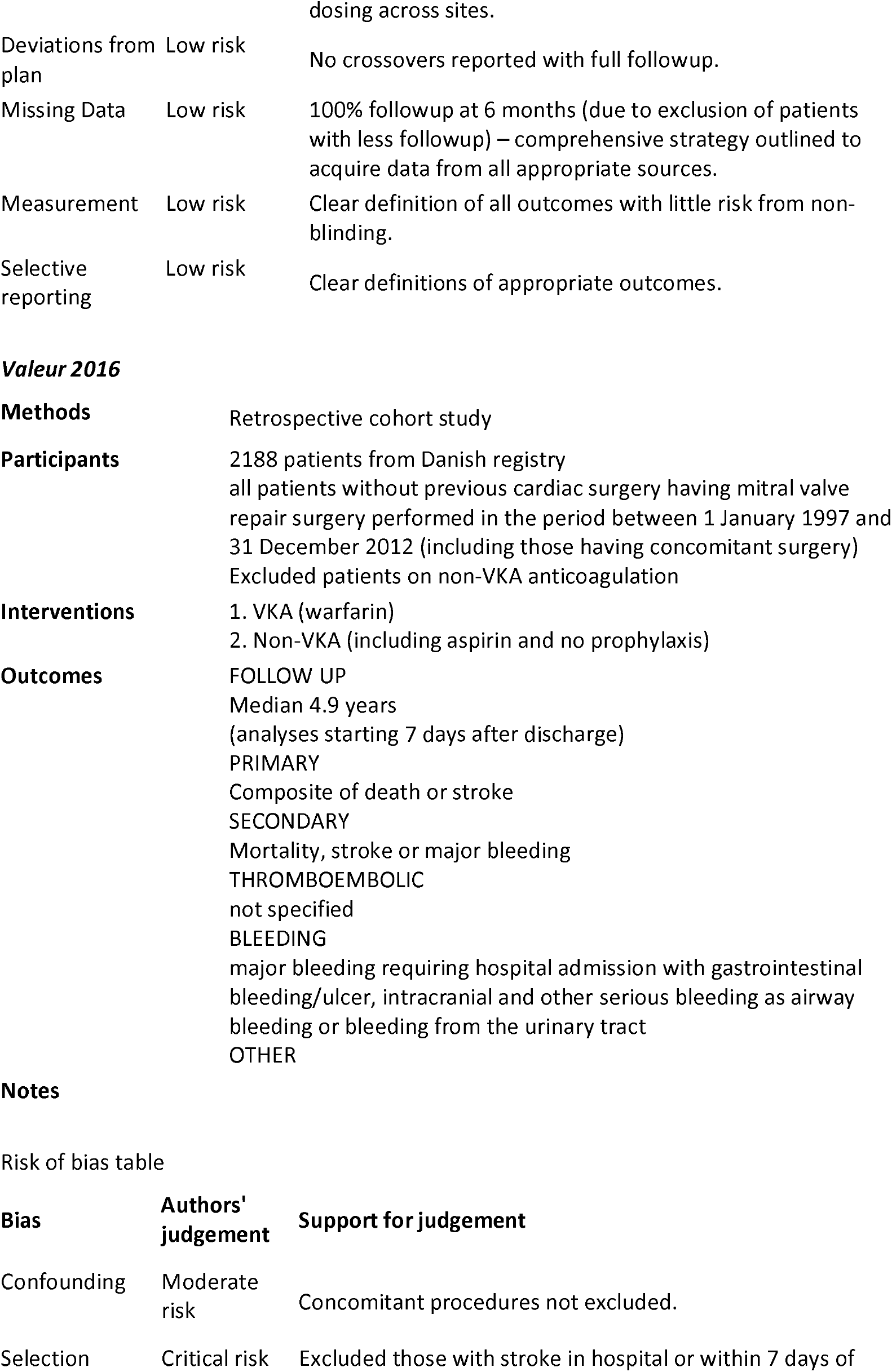
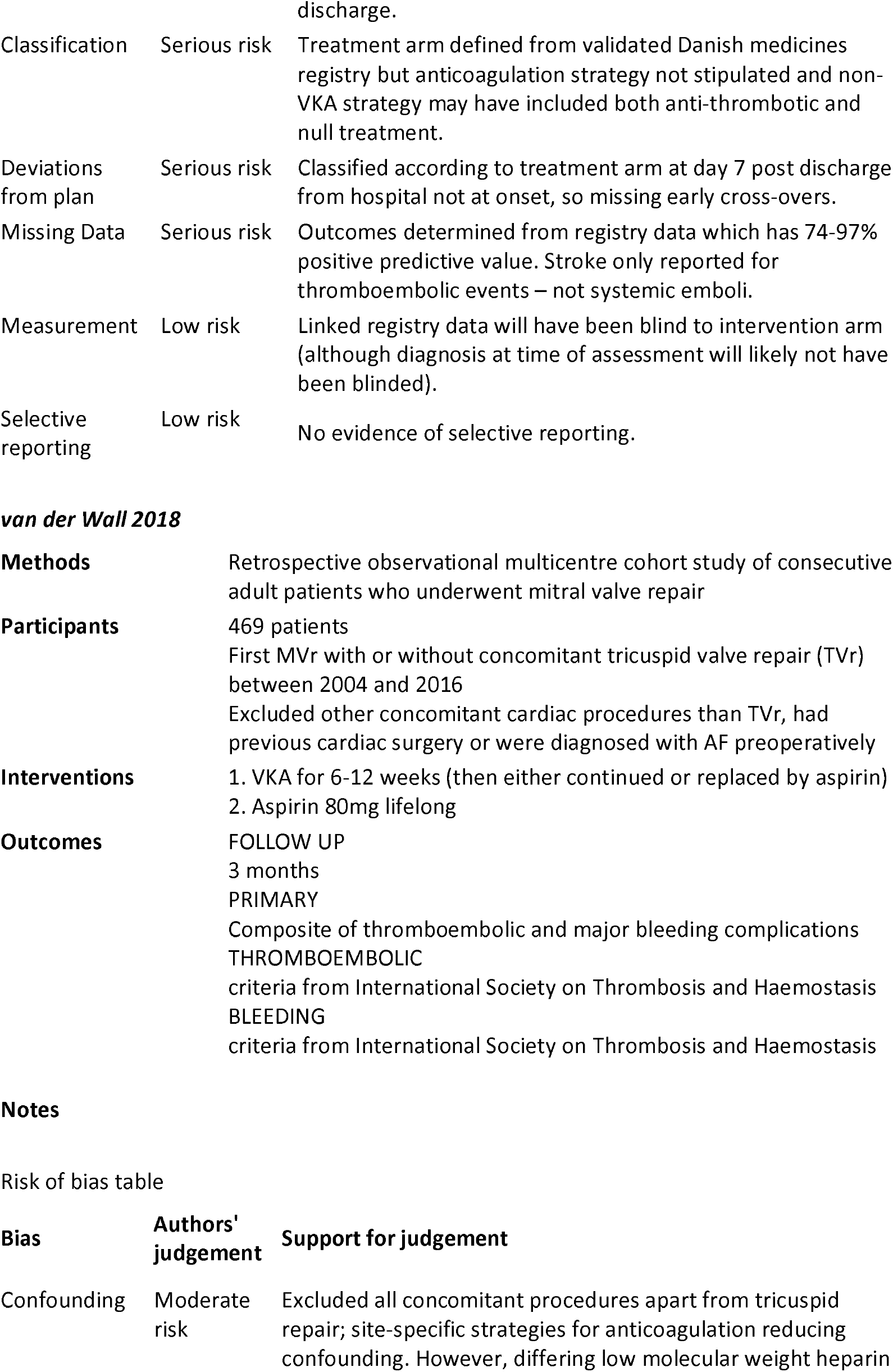
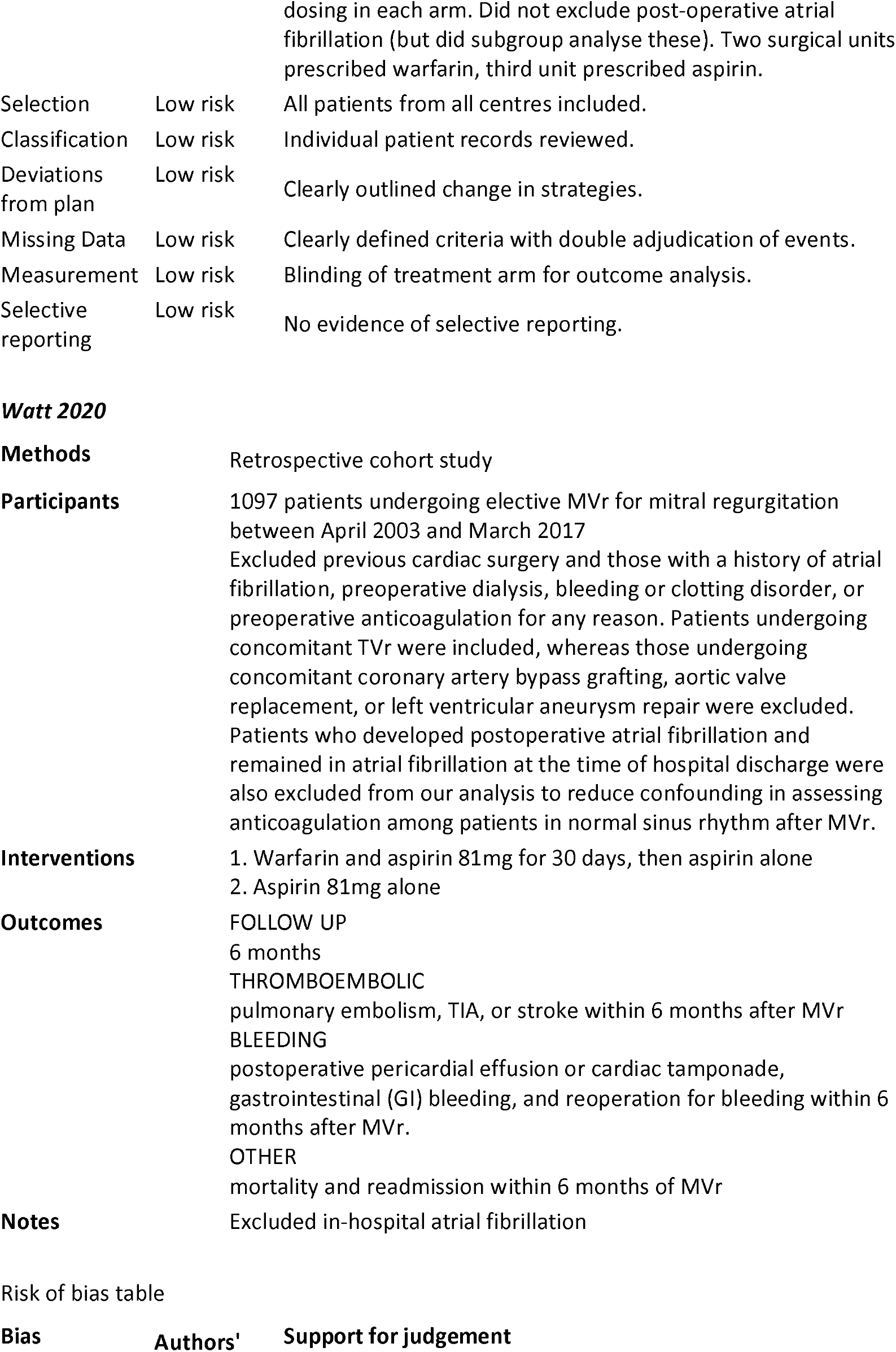
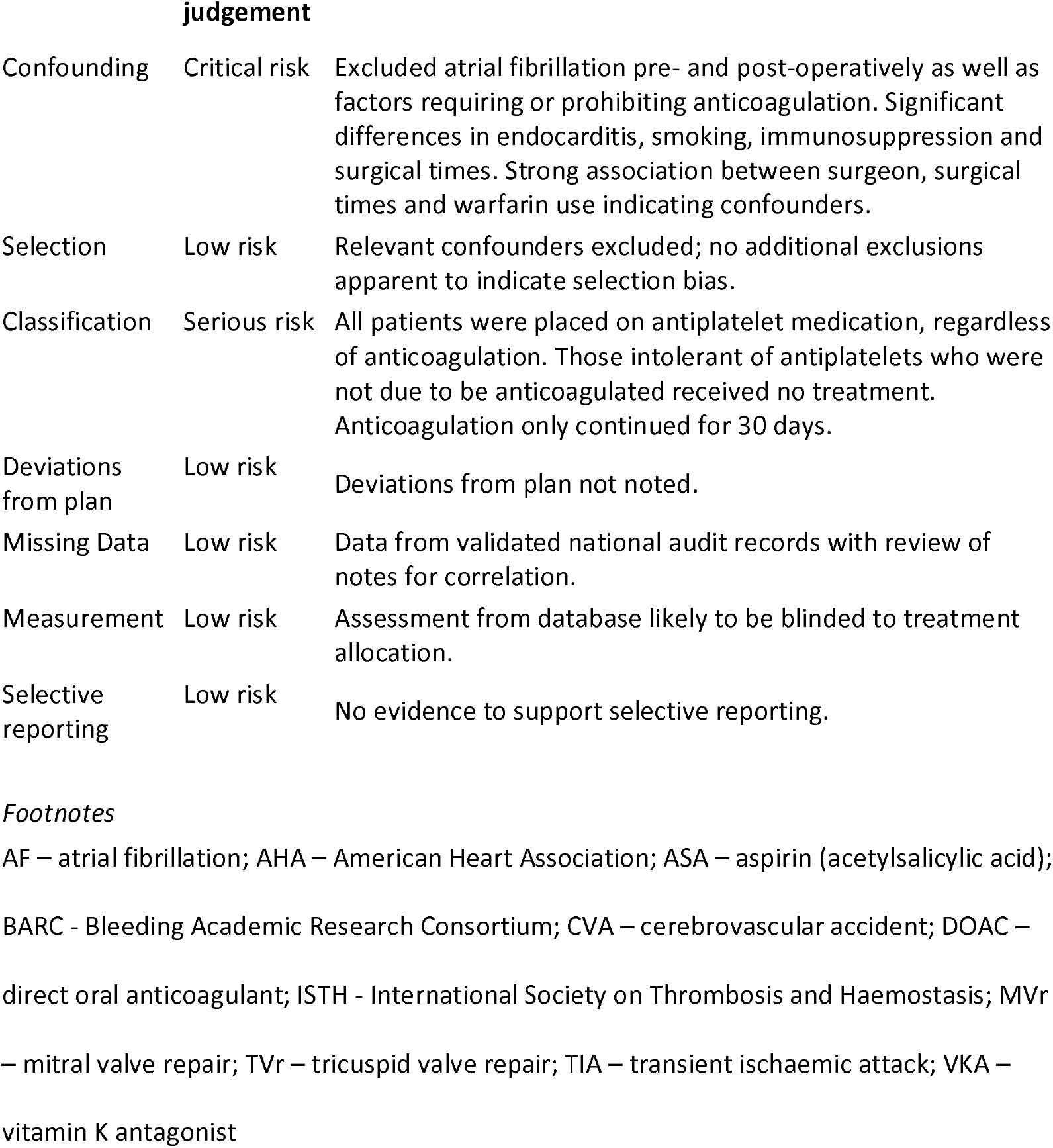

